# Changes in Cause-of-Death Attribution During the Covid-19 Pandemic: Association with Hospital Quality Metrics and Implications for Future Research

**DOI:** 10.1101/2020.07.25.20162198

**Authors:** Kathleen A. Fairman, Kellie J. Goodlet, James D. Rucker

**Affiliations:** Midwestern University College of Pharmacy-Glendale, Glendale, Arizona; Department of Pharmacy Practice, Midwestern University College of Pharmacy-Glendale, Glendale, Arizona; Kathleen Fairman LTD, Phoenix, AZ

## Abstract

**Background:** Severe acute respiratory syndrome coronavirus 2 (SARS-CoV-2) is often comorbid with conditions subject to quality metrics (QM) used for hospital performance assessment and rate-setting. Although diagnostic coding change in response to financial incentives is well documented, no study has examined the association of QM with SARS-CoV-2 cause-of-death attribution (CODA). Calculations of excess all-cause deaths overlook the importance of accurate CODA and of distinguishing policy-related from virus-related mortality.

**Objective:** Examine CODA, overall and for QM and non-QM diagnoses, in 3 pandemic periods: awareness (January 19-March 14), height (March 15-May 16), and late (May 17-June 20).

**Methods:** Retrospective analysis of publicly available national weekly COD data, adjusted for population growth and reporting lags, October 2014-June 20, 2020. CODA in 5 pre-pandemic influenza seasons was compared with 2019-20. Suitability of the data to distinguish policy-related from virus-related effects was assessed.

**Results:** Following federal guidance permitting SARS-CoV-2 CODA without laboratory testing, mortality from the QM diagnoses cancer and chronic lower respiratory disease declined steadily relative to prior-season means, reaching 4.4% less and 12.1% less, respectively, in late pandemic. Deaths for non-QM diagnoses increased, by 21.0% for Alzheimer’s disease and 29.0% for diabetes during pandemic height. Increases in competing CODs over historical experience, suggesting SARS-CoV-2 underreporting, more than offset declines during pandemic height. However, in the late-pandemic period, declines slightly numerically exceeded increases, suggesting SARS-CoV-2 overreporting. In pandemic-height and late-pandemic periods, respectively, only 83.5% and 69.7% of increases in all-cause deaths were explained by changes in the reported CODs, including SARS-CoV-2, preventing assessment of policy-related mortality or of factors contributing to increased all-cause deaths.

**Conclusions:** Substitution of SARS-CoV-2 for competing CODs may have occurred, particularly for QM diagnoses and late in the pandemic. Continued monitoring of these trends, qualitative research on pandemic CODA, and the addition of place-of-death data and psychiatric CODs to the file would facilitate assessment of policy-related and virus-related effects on mortality.

Ascertainment of the number of deaths from severe acute respiratory syndrome coronavirus 2 (SARS-CoV-2) is foundational to understanding the severity, scope, and spread of the infection. Despite its importance, estimation of SARS-CoV-2 deaths is challenging because advanced age, genetic polymorphisms, and obesity-related comorbidities that predispose to inflammatory states increase the likelihood of dysregulated immunological function, severe respiratory distress, and mortality from infectious respiratory illness.^1,2^ These host factors represent competing potential causes of death (COD). For example, 98.8% of Italy’s SARS-CoV-2 deaths occurred in persons with ≥1 comorbidity, 48.6% with ≥3 comorbidities, and median decedent age was 80 years.^3^ Similarly, of U.S. SARS-CoV-2 deaths reported as of May 28, 2020, 93% involved other CODs (mean 2.5 additional causes), and 60% occurred in persons aged ≥75 years.^4^ This pattern of multiple contributing CODs is common in respiratory infection-related mortality.^5^

In death certificate issuance during the pandemic, methods to account for this pattern varied, as no single standard for SARS-CoV-2-attributable death exists. In Italy, all deaths in patients testing positive for SARS-CoV-2 were attributed to the infection despite high prevalence rates for comorbid conditions, measured in early deaths: ischemic heart disease (30%), diabetes (36%), cancer (20%), and atrial fibrillation (25%).^6^ The U.S. National Center for Health Statistics (NCHS) issued death-certification guidance on March 4, 2020, indicating that SARS-CoV-2 should be reported if “the disease caused or is assumed to have caused or contributed to death.”^7^ Follow-up guidance issued on April 3 indicated that it was “acceptable to report COVID-19 on a death certificate without [laboratory test] confirmation” if circumstances indicating likely infection were “compelling within a reasonable degree of certainty.”^8^

This nonspecific guidance should be interpreted in light of previous research findings that COD attribution (CODA) errors are common on death certificates, particularly in infectious disease and septic shock.^9,10^ In one survey of New York City (NYC) resident physicians in 2010, 49% indicated they had knowingly reported an inaccurate COD on one or more certificates, often (54%) at the behest of hospital staff, and 70% reported they had at least once been unable to report septic shock “as an accepted cause of death” and had been “forced to list an alternate cause.”^9^ In an audit of NYC data from 2010-2014, 67% of pneumonia death certificates contained ≥1 error, compared with 46% for cancer and 32% for diabetes.^10^

Such CODA ambiguities are often addressed by calculating “excess deaths,” defined as all-cause deaths exceeding those projected from historical experience.^11^ This method, recently used to estimate that official tallies of SARS-CoV-2 deaths represented only about 66%-78% of the disease’s true mortality impact,^12,13^ is potentially advantageous in estimating SARS-CoV-2 impact by accounting for deaths that may not have been explicitly coded as infection-related.^14^ Examples include deaths from cardiac events to which undetected SARS-CoV-2 may have contributed^15^ or out-of-hospital deaths occurring without medical care because of health-system overcrowding.^16^ Despite these advantages, the method is compromised by 3 considerations when applied to SARS-CoV-2 CODA, which should be quantified to inform future policy.

First, the method should distinguish natural from societal causes to account for possible consequences of policy decisions and fears that, although prompted by anticipated effects of SARS-CoV-2, were not direct or inevitable viral sequelae. Examples include suicides from stay-at-home order-related labor market contraction^17^ and social isolation,^18^ increases in domestic violence,^19^ overdoses due to interruptions in substance use disorder treatment,^20^ and delays in emergency care for life-threatening conditions^21–23^ in geographic areas where health-system overcrowding was expected but not realized.^24,25^ To promote evidence-based public health policy, population-level disease-mitigation strategies that go beyond traditional practices of isolating the sick and quarantining those exposed to disease merit empirical investigation.^26,27^

Second, the method should reflect the effects that financial incentives around hospital quality metrics (QM), which are commonly associated with provider coding practices, may have on CODA.^28–30^ For example, in United Kingdom hospitals, increases in coding for palliative-care admissions produced a severity-adjusted mortality-rate decline of 50% over the 5-year period ending in 2009, while the crude death rate remained unchanged.^31^ Although we are not aware of studies linking QMs to CODA, it is known that CODA errors are more likely to occur in hospitals than elsewhere,^32^ with an 85% error rate reported in comparisons of death certificates with autopsy findings at one regional academic institution.^33^ The potential effect of financial incentives on CODA is particularly important for SARS-CoV-2 because several competing CODs, including chronic lower respiratory disease (CLRD), acute myocardial infarction, heart failure, pneumonia, and stroke, are included in Medicare 30-day mortality measures used to calculate prospective payment rates.^34^ All but one of these (CLRD) is included in Agency for Healthcare Research and Quality inpatient quality indicators.^35^ Sepsis and cancer, other competing causes of death, are also the target of QM.^36–38^ Although not affecting all-cause death counts, the incentive to substitute SARS-CoV-2 for another COD could affect the accuracy of the SARS-CoV-2-attributed count.

Third, the method should account for baseline life expectancies among those whose deaths were reported as caused by SARS-CoV-2. For example, at age 80 years, the 1-year probability of death is 5.8% for males and 4.3% for females, higher in those with cardiovascular comorbidities.^39,40^ In that age group, the population-level risk of a SARS-CoV-2 death in New York City, a pandemic epicenter, was 1.5% in about 3 pandemic months through June 17, 2020.^41^ Thus, deaths from competing CODs would be expected to decline late in the pandemic and in subsequent months. From a policy perspective, quantifying this effect is consistent with the quality-adjusted life year approach in evidence-based medicine, which considers future life expectancy in assessing the effects of disease and disease-mitigation interventions.^42^

To permit assessments of SARS-CoV-2-related mortality, publicly available NCHS data include weekly aggregated totals for all-cause deaths, natural-cause deaths, and selected categories of CODs, reported as final data for 2014-2018 and provisional data for 2019-2020.^4^ These data, which are updated weekly, have important limitations. First, International Classification of Diseases (ICD)-10 diagnosis codes are grouped into broad categories, rather than the individual ICD-10 codes available in full COD files (Appendix 1). Second, only 11 selected diagnostic categories are reported. Third, although 63% of deaths are reported within 10 days, reporting lags vary by state.^4^ Reporting delays for injurious deaths are greater because they require investigation (e.g., forensic toxicology).^43^ Pending investigation, these deaths are often assigned ICD-10 code R99, “ill-defined and unknown cause of mortality.”^43^

In this exploratory study, we used these files to provide preliminary evidence on the following: (1) change in CODA compared with historical experience; (2) association of CODA with QM; and (3) suitability of the files to distinguish policy-related from virus-related effects. All analyses were adjusted for population and reporting lags and based on comparisons of 2020 with equivalent weeks in the 5 most recent years. We hypothesized that if substitution of SARS-CoV-2 for alternative CODs occurred, death counts for competing diagnoses would decline relative to historical experience during the pandemic, especially after issuance of the NCHS death-certification guidance; these declines would be greater for QM than for other conditions; and they would accelerate late in the pandemic as earlier SARS-CoV-2 deaths offset later deaths from competing causes.

## Methods

### Data

Publicly available COD data for October 2014 through June 20, 2020, were downloaded from the NCHS website on July 10, 2020. The outcome was reported underlying COD (UCOD), defined as “the disease or injury which initiated the train of morbid events leading directly to death.”^44^ Three states were excluded because of incomplete reporting in 2020 (Connecticut and North Carolina) or in 2015 (Wisconsin) by subtracting their death counts and population counts from U.S. figures. Raw death counts were adjusted to July 2019 population using annual census data (Appendix 2).^45^

Reporting lag adjustment was made by calculating percentage changes in death counts, overall and by ICD category, for each of 7 weekly updates from May 27 to July 8, 2020, as a function of the number of weeks from the reported week to the update week (e.g., deaths in week ending May 9, updated on May 20 = 1.6-weeks). For each of 16 weeks prior to the July 8 update, we used mean percentage change values as inflationary factors to calculate compounded adjustment multipliers (Appendix 3).

### Statistical Analyses

Time series analyses were performed using standard methods, beginning with descriptive analysis.^46,47^ We first plotted mortality curves over time, then compared pandemic periods with mean values for the same weeks in prior seasons. Absolute and percentage changes in counts of deaths, comparing 2020 with 5-year means, were calculated for 3 pandemic periods: pandemic awareness beginning January 19, based on media-report analysis of news coverage volume;^48^ pandemic height beginning March 15, the approximate first date of >1,000 U.S. cases;^49^ and late pandemic, beginning May 17. These calculations were performed separately for all-cause deaths; for each ICD category; and for grouped categories based on potential QM inclusion, defined as percentage of deaths in the category that were QMs in the most recent full COD file (2018; Appendix 1): all or nearly all QM (cancer, CLRD, septicemia; ≥97% of deaths); some QM (cerebrovascular disease [CBVD], heart disease, influenza/pneumonia; 56%-81%); and no QM (Alzheimer’s disease, diabetes, kidney disease, other acute respiratory illness).

For all-cause deaths and separately for each category, we fit Poisson regression models with Fourier terms (sine-cosine pairs) to control for annual and semiannual seasonality. Models were calibrated through the first 5 years (October 2014-September 2019), and the resulting equations applied to the 2019-20 influenza season to generate predicted values. Prediction confidence ranges were estimated using Wald confidence interval equation parameters. Because all models except cancer and deaths not elsewhere classified (NEC) displayed classic first-order autocorrelation, with gradual declines in autocorrelation function and precipitous declines in partial autocorrelation function correlograms after lag 1,^50^ we added first-order autoregression (AR1) terms to the models. When the resulting models produced the unexpected and seemingly erroneous result of only 21,833 excess all-cause deaths during pandemic height, we performed additional modeling excluding the AR1 terms, thereby permitting autocorrelated errors. All analyses were performed using IBM SPSS v25.0 (Armonk, NY) with a priori alpha=0.05.

## Results

Of approximately 15 million population-adjusted all-cause deaths reported in the 47 study states during the 5.8-year study period, 10.7 million (71%) were reported in one of the ICD categories included in the COD file, of which 400,000 were reported as a death NEC, including the ICD code R99. Counts formed a seasonal annual pattern, with defined peaks and troughs at similar times each year (Appendix 4). Exceptions in 2020 included a higher and later peak in all-cause deaths in April; a later peak for heart disease deaths, also in April; a marked increase in NEC deaths; and declines in deaths from cancer and CLRD. The 1-week maximum peak in all-cause deaths increased every 2-3 years, with a greater increase in 2020 than in previous years: 58,502 in 2015, 62,979 in 2018, and 73,367 in 2020. Deaths in the final study week, ending June 20, 2020, appeared to decline sharply. Although declines of this magnitude were not unusual in 2020, we added a sensitivity analysis excluding the final week to address possible residual underreporting.

Comparisons of 2020 values with prior-year population-adjusted means suggested a peak in all-cause deaths beginning at pandemic start and returning to near mean values approximately 12 weeks later, corresponding closely to a similar peak in total acute respiratory deaths, including influenza/pneumonia, other acute respiratory illness, and SARS-CoV-2 (Figure 1, Panel 1; Table 1). Influenza/pneumonia deaths peaked later in the season in 2020 than in prior years, despite similar magnitudes. NEC deaths including R99 increased rapidly beginning in January, exceeding 5-year means by 53% during the pandemic awareness period and by >100% during and after pandemic height.

**Table 1.**
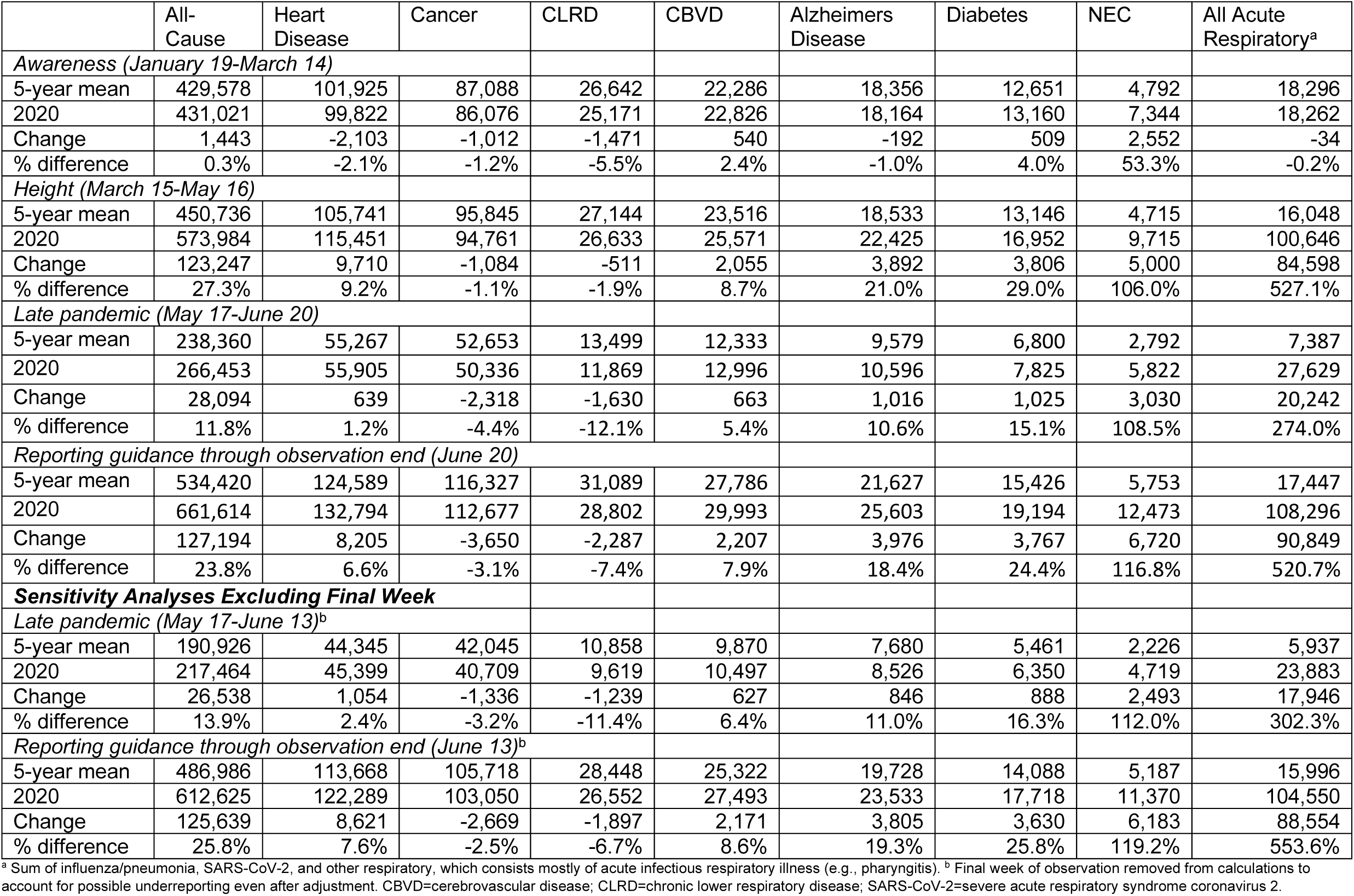
Population-Adjusted and Reporting Lag-Adjusted Mortality Counts, 5-Year Means and 2020 Totals, All-Cause and Selected Causes of Death, by Pandemic Time Period

**Figure 1.**
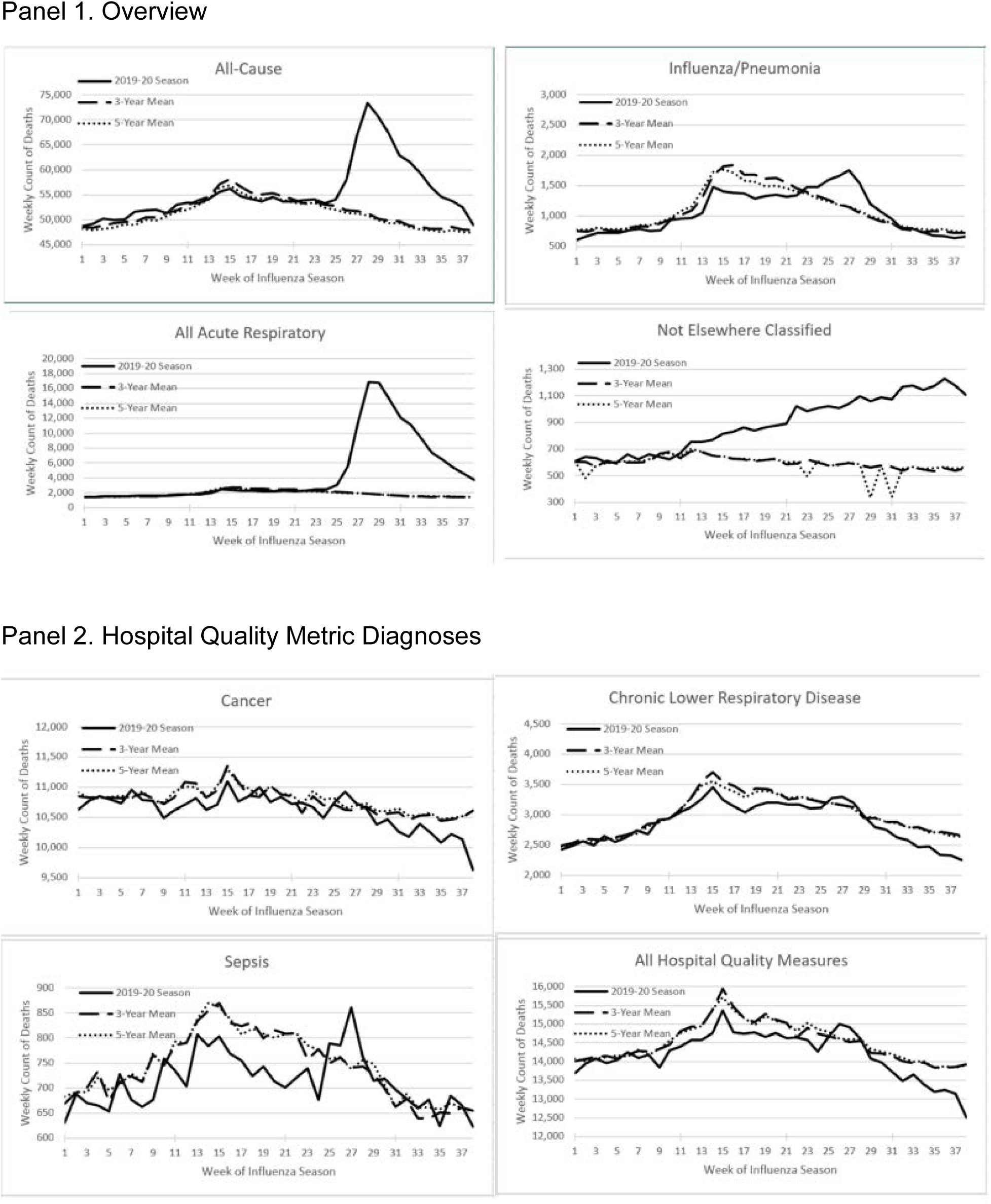
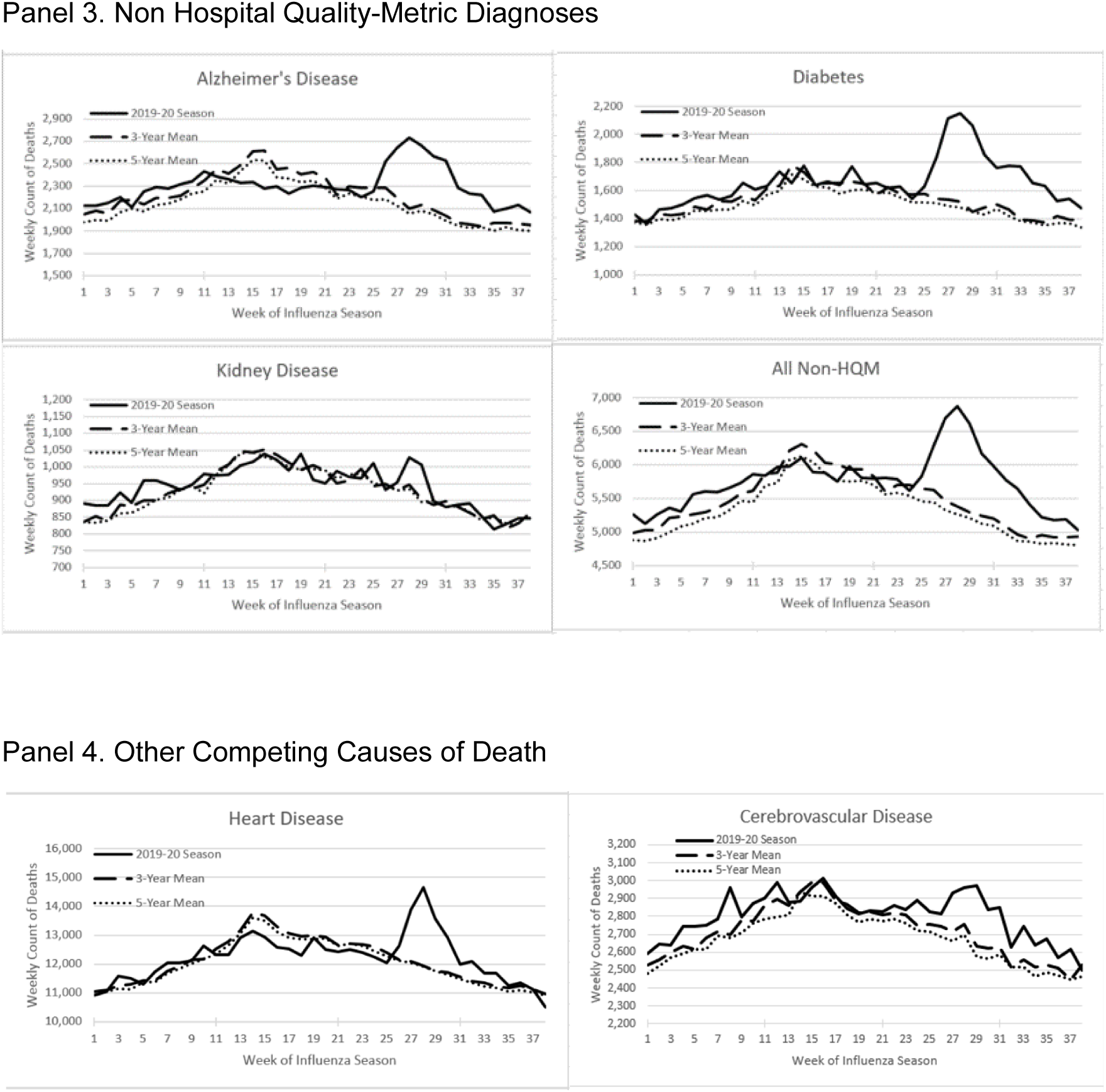
Population-Adjusted and Lag-Adjusted Weekly Death Counts, by Week of Season, 5-Year and 3-Year Prior Season Means and 2019-20 Notes: Season week 1 begins September 29-October 5, depending on year. Five-year means are seasons 2014-15 through 2018-19, and 3-year means are 2016-17 through 2018-19. Season week 17 (January 19) is the start of pandemic awareness (beginning of intense media coverage of the pandemic).^48^ Season week 25 (March 15) is the approximate start of the pandemic height period. Season week 28 (April 5) corresponds to the issuance of NCHS guidance stating laboratory testing was recommended but not required to name SARS-CoV-2 as the underlying cause of death. Season week 34 (May 17, 2020) is the start of the late pandemic period. Acute respiratory (Panel 1) is the sum of SARS-CoV-2, influenza/pneumonia, and other respiratory illnesses, nearly all of which are acute (e.g., pharyngitis). Not elsewhere classified (Panel 1) includes R99, the code used for non natural-cause deaths pending forensic investigation. QM diagnoses include cancer, chronic lower respiratory disease, and septicemia. Non-QM diagnoses include Alzheimer’s disease, diabetes, kidney disease, and other respiratory illnesses. QM=quality metric; SARS-CoV-2=severe acute respiratory syndrome coronavirus 2.

Following issuance of NCHS reporting guidance in the first week of April 2020, deaths for 2 of 3 QMs, cancer and CLRD, but not for sepsis, steadily declined relative to prior year means (Figure 1, Panel 2; Table 1). Although these declines were initially small (1%-2%) and within the fluctuation observed during pandemic awareness, late-pandemic cancer deaths were 4.4% less and CLRD deaths 12.1% less than in prior years (3.2% and 11.4%, respectively, in the sensitivity analysis excluding the final study week). Measured from guidance issuance to study end, these changes represented declines of 5,937 deaths (4,566 in sensitivity analysis) for these 2 QMs, compared with prior years.

For non-QM diagnoses, these declines did not occur; instead, deaths increased beginning at pandemic start, including increases of 21.0% for Alzheimer’s disease (3,892 deaths) and 29.0% for diabetes (3,806 deaths) during pandemic height (Figure 1, Panel 3; Table 1). Deaths from other competing CODs, for which some but not all diagnoses are QMs, increased by smaller amounts than non-QM diagnoses (CBVD by 8.7%, heart disease by 9.2%; Figure 1, Panel 4; Table 1).

During pandemic height, increases in the non-QM or partial-QM diagnoses more than offset declines in the QM diagnoses (net effect of +21,714 deaths excluding respiratory illness; not shown in Table 1). However, of the total increase of 123,247 all-cause deaths compared with prior-year mean, only 102,956 deaths (83.5%) were explained by changes in the specific CODs included in the NCHS files, leaving 16.5% of the increase in all-cause deaths unexplained.

By the late pandemic period, 2 additional competing CODs had declined compared with historical experience: influenza/pneumonia (−11.6%) and other acute respiratory illnesses (−3.8%), whereas other competing CODs were relatively unchanged (+/−1%, heart disease, septicemia, and kidney disease) or remained elevated (Alzheimer’s disease, 10.6%; diabetes 15.1%; CBVD 5.4%; not all percentages shown in Table 1). In this time period, the net effect of competing CODs was a small decline (−1,232 deaths), and 30.3% of the increase in all-cause deaths was unexplained by specific CODs.

Time series models using AR1 terms fit the data well during the calibration period (October 2014-September 2019; Appendix 5). However, when equations were applied to the 2019-20 influenza season, results were highly sensitive to modeling method (Appendix 6). When modeling was performed using AR1 terms to account for first-order autocorrelation, results suggested only 21,833 excess all-cause deaths during pandemic height. When the nonautocorrelation assumption was intentionally violated by excluding AR1 terms, that estimate increased to 87,943 during pandemic height and 42,964 in late pandemic, for a total of 130,907. Despite this major discrepancy, both models showed greater magnitudes in CODA for non-QM than for QM conditions, as well as less-than-expected death counts for QMs in the late pandemic period.

## Discussion

In this exploratory analysis of NCHS COD files, we found evidence suggesting SARS-CoV-2 may have been substituted for 2 of 3 competing CODs associated with QMs after the NCHS published guidance specifying no laboratory testing requirement for presumptive SARS-CoV-2 UCOD. We also found decreases in several competing CODs relative to prior year means, offset by marked increases in reported deaths from other conditions during pandemic height. Although validating assertions that for patients with certain comorbidities, SARS-CoV-2 may have been underreported as a UCOD,^51^ our findings suggest these assertions are oversimplified because of overreporting of SARS-CoV-2 for those with other comorbidities including the second and third top causes of natural death in the United States, cancer and CLRD.^52^ Despite concerns about underreporting of SARS-CoV-2 on death certificates,^53^ overreporting began to slightly numerically exceed underreporting by the late pandemic period.

Our methods were generally similar to those of the previously reported studies of excess deaths,^12,13^ although our method of adjustment for reporting lags was simpler and our study period was longer. The primary strength of the present study was our examination of specific CODs, rather than all-cause deaths alone, because from a policy perspective, the quality of mortality data is important.^53^ Moreover, we assessed the suitability of the files to distinguish policy-related from virus-related mortality.

Information of this type should be viewed as essential in considering responses to future pandemics.

Like previous investigators of excess deaths,^12^ we found large (122,300 in previous analysis, 130,907 in our analysis) increases in all-cause deaths compared with prior years in Poisson regression models adjusted for seasonality using Fourier terms, but only when our model, like the previous model, included no term to control for autocorrelation. In the model including an AR1 term, we found only a small (21,833) increase in all-cause deaths. It is possible the AR1 model better accounted for the gradually increasing peak in all-cause mortality that occurred biannually or triannually from 2014-15 to 2019-20. However, it is also possible that the model is misleading because of excessively data-driven specification.^46^ Therefore, we view the time series models as inconclusive, basing our interpretation primarily on descriptive analyses of the population-adjusted data.

Our finding of potential underreporting of QM-associated deaths compared with prior years is consistent with our a priori hypothesis, which was based on previous research documenting institutional sensitivity of coding practices to policy and financial incentives.^28–31^ The present study adds to the body of literature on this topic by associating these effects with death certification. This finding suggests a need for qualitative assessment of how CODAs were made during the pandemic, perhaps conducted by the Office of Inspector General, as QM data are used to set Medicare prospective payment rates.^34^ A potential pragmatic contribution of that research is a specific CODA algorithm to be used in future pandemics, facilitating valid and reliable reporting across sites (e.g., hospitals versus nursing homes) and geographic regions. Important limitations of this exploratory work should be noted. Foremost, findings of this study should be reassessed using data from later weeks to verify or refute the preliminary trends described here. Additional limitations suggest improvements that might be made to the COD files pending availability of the full, final COD data. These full COD data, which are currently available only through 2018, likely will not be posted until 2022.

First, a large number of deaths had no COD or a nonspecific COD recorded in the file. Compared with prior years, NEC deaths more than doubled in a progression that began early in 2020, as pandemic awareness was growing but before SARS-CoV-2 would have been expected to have any biological effect. Moreover, of increases in all-cause death during the pandemic, 16.5% during pandemic height and 30.3% in the late-pandemic period could not be linked to change in the specific CODs included in the file. Based on national evidence of increases in prescriptions for psychotropic drugs beginning on February 16, 2020,^54^ reported tripling of depression or anxiety comparing April and May 2019 to 2020,^55^ and unprecedented media coverage of SARS-CoV-2 compared with other pandemics,^48^ measurement of the mental health effects of growing pandemic awareness and stay-at-home orders on mortality would likely be helpful. This measurement is not possible with the NCHS COD files as currently constructed but could be facilitated with the addition of a new category for psychiatric UCODs (i.e., ICD-10 category F, excluding intellectual disabilities and developmental disorders). Similarly, disaggregation of R99, which is used pending forensic investigation of injurious deaths,^43^ from the rest of the NEC category would facilitate assessments of potential deaths from suicide or overdose.

Second, we imputed possible effects of financial incentives on CODA but did not measure incentives directly. To support or refute our findings, the addition of place of death to the COD files would allow for assessment of whether the QM associations we observed were greater in hospital than in nonhospital settings. Additionally, this change would facilitate assessment of changes in out-of-hospital deaths from heart disease or cerebrovascular disease,^21–23^ informing the question of whether stay-at-home orders exacerbated deaths from these causes in geographic areas where hospital bed supply was not limited during the pandemic.^24–25^

Third, neither the study outcome measure nor recommended changes to the COD files would account for long-term physiological effects of SARS-CoV-2, which are the subject of an ongoing cohort study;^56^ of suboptimal policy decisions that could have increased SARS-CoV-2 deaths, (e.g., mandatory posthospitalization release of infected patients to nursing facilities);^57^ or of long-term effects of stay-at-home order-related declines in preventive care (e.g., pediatric immunizations).^58^

Finally, it should be noted that although recent comparisons of infection-fatality and case-fatality rates for influenza and SARS-CoV-2 have adopted an implicit assumption that the respective infections’ numerators are equivalent,^59^ the 2 estimates are based on markedly different methods that make direct comparison problematic. National estimates of influenza hospitalizations and deaths are based on a statistical model incorporating ambulatory and inpatient data, in addition to the raw death certificate counts reported here.^60^ In contrast, SARS-CoV-2 death counts rely solely on the judgment of the reporter and are, at present, not clinically validated. Pending assessment of the reliability and validity of SARS-CoV-2 death reporting, policy making death-certificate counts should be circumspect, recognizing both the possible vulnerability of the SARS-CoV-2 data to financial incentives and detection bias and the absence of important COD information from the currently available NCHS files.

## Conclusion

SARS-CoV-2 deaths appear underreported with some comorbidities, such as Alzheimer’s disease and diabetes, and overreported with others including cancer and CLRD, the second and third leading causes of natural death in the United States.

Possible overreporting slightly exceeded underreporting for competing CODs beginning late in the pandemic and merits further investigation. Relatively small additions to available NCHS COD data—including place of death, disaggregation of R99 from other NEC deaths, and psychiatric UCODs—could facilitate preliminary analyses of important questions about direct effects of SARS-CoV-2, as well as the effects of stay-at-home orders and media-driven pandemic awareness, on mortality in the pandemic period.

**Appendix 1.**
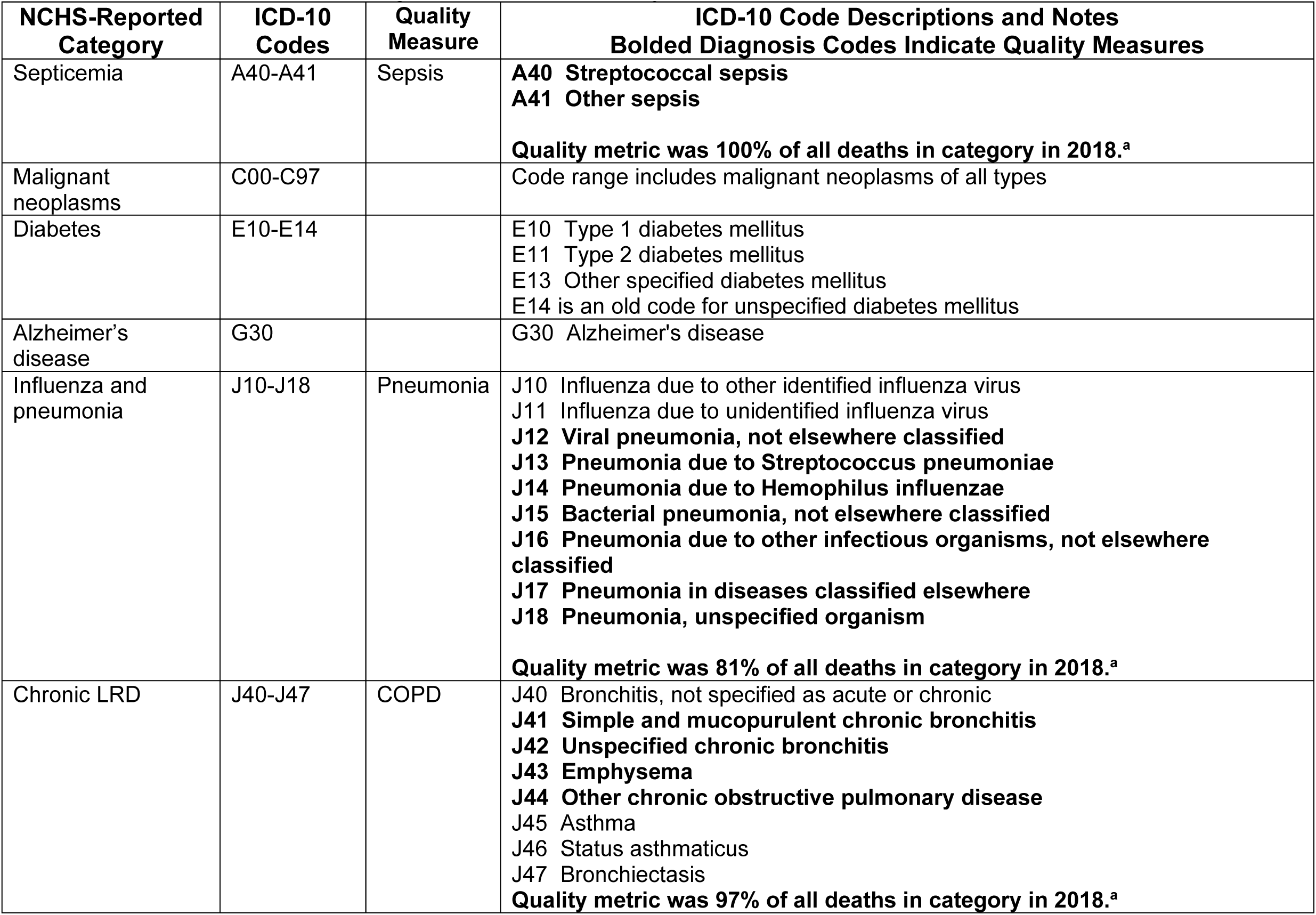
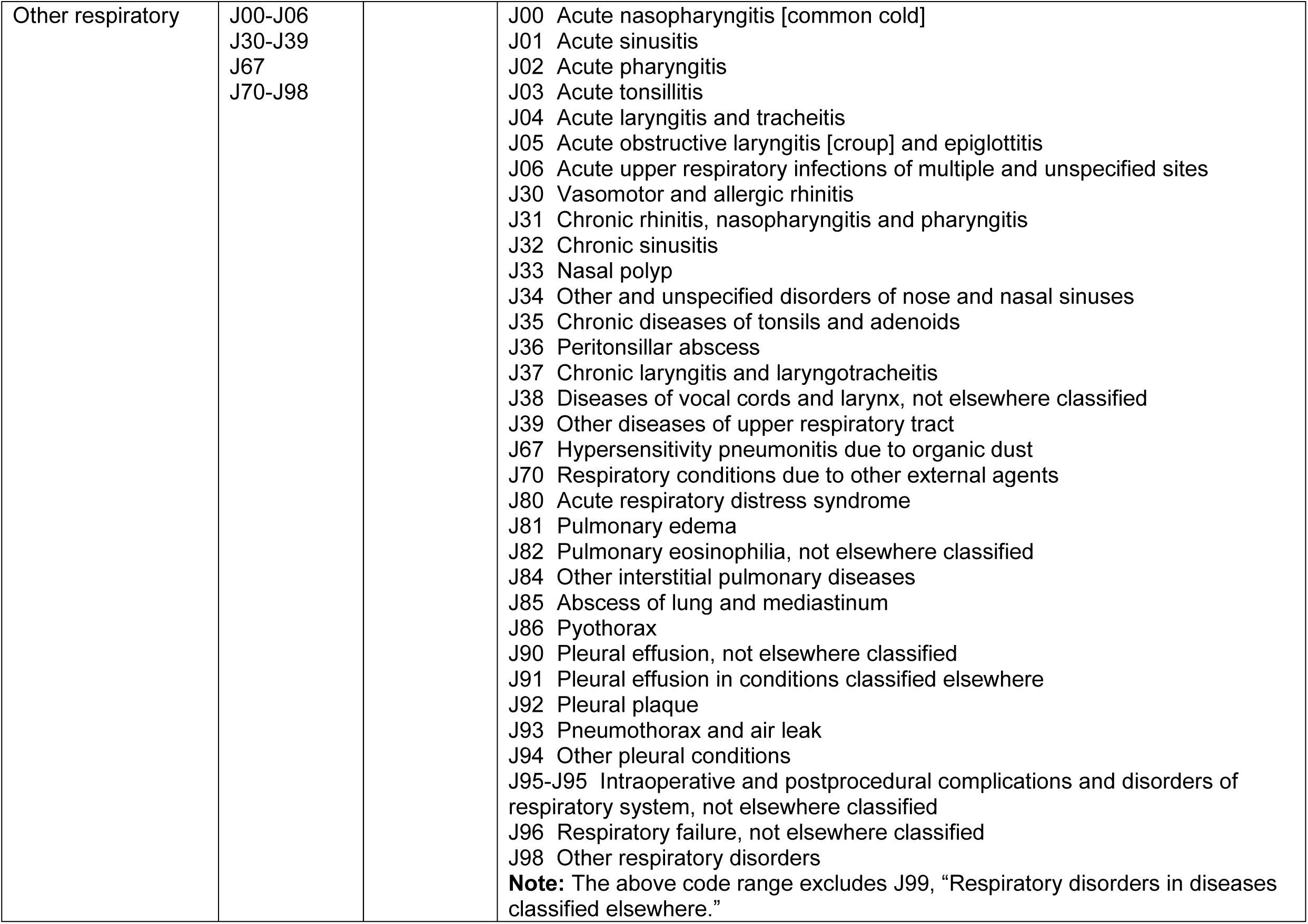
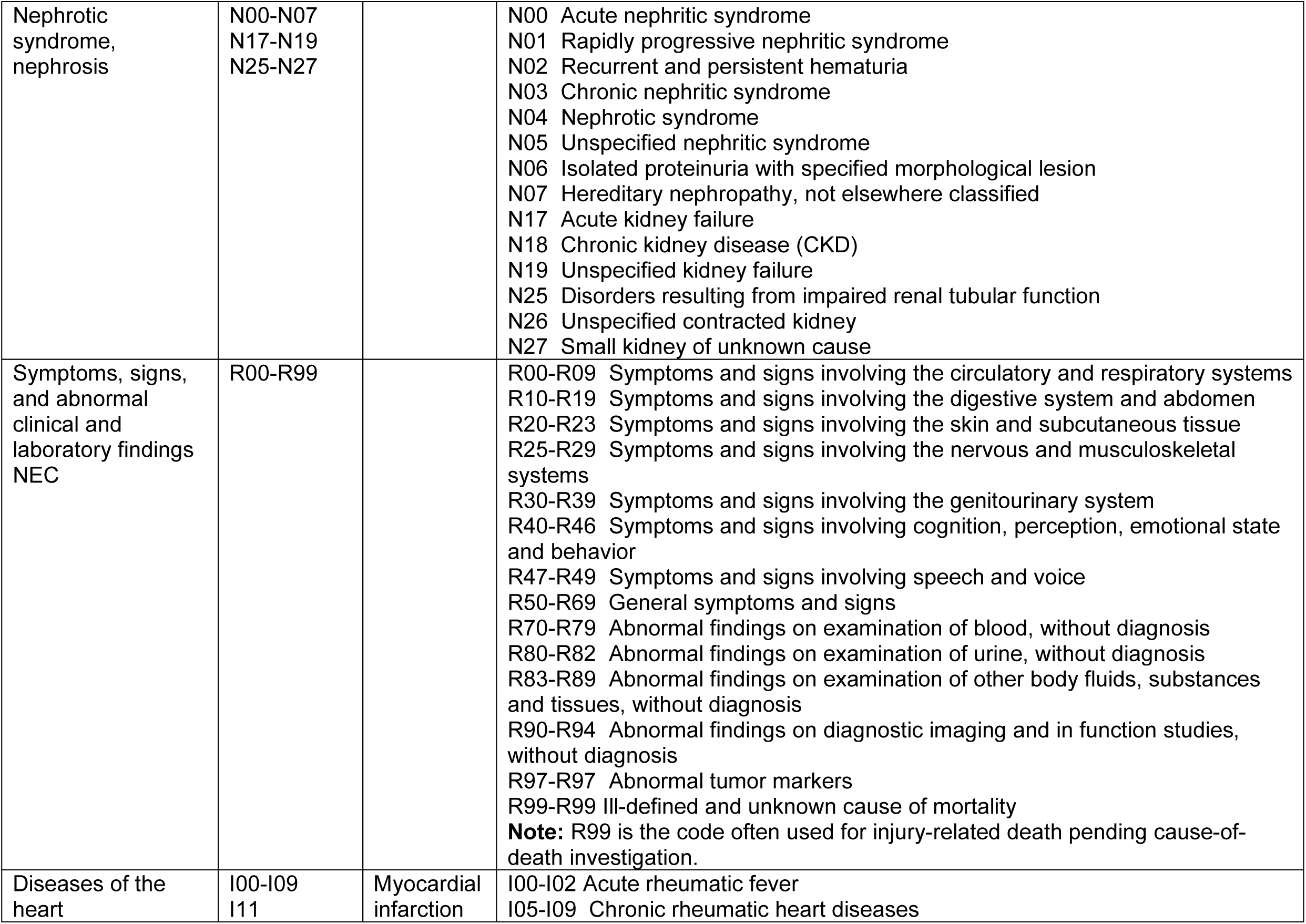
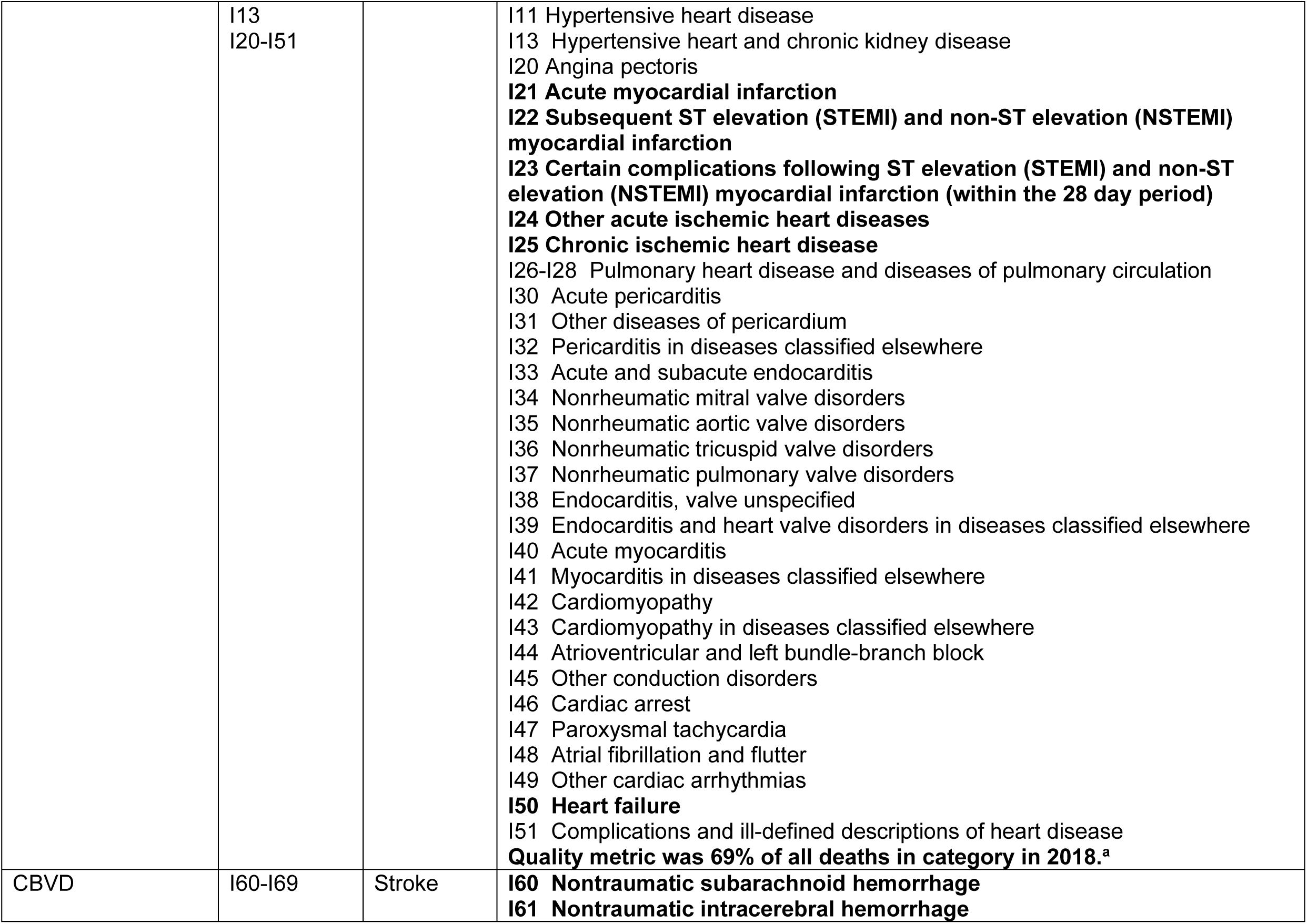
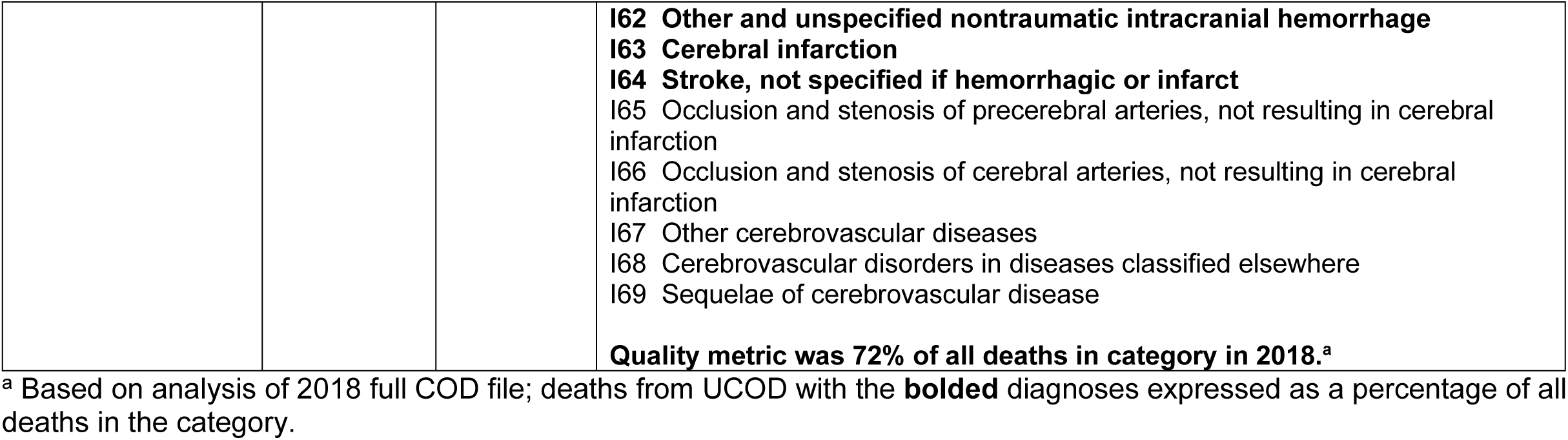
ICD-10 Code Groupings in NCHS Preliminary Cause-of-Death Files and Specific Code Descriptions

**Appendix 2.**
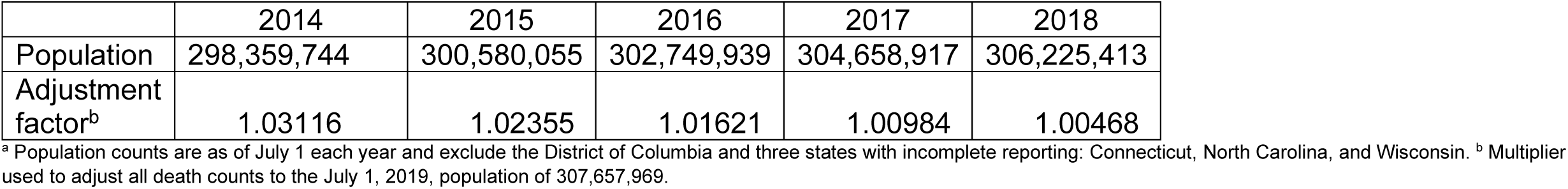
Populations and Adjustment Factors^a^

|  | 2014 | 2015 | 2016 | 2017 | 2018 |
| --- | --- | --- | --- | --- | --- |
| Population | 298,359,744 | 300,580,055 | 302,749,939 | 304,658,917 | 306,225,413 |
| Adjustment factor<sup>b</sup> | 1.03116 | 1.02355 | 1.01621 | 1.00984 | 1.00468 |

<sup>a</sup> Population counts are as of July 1 each year and exclude the District of Columbia and three states with incomplete reporting: Connecticut, North Carolina, and Wisconsin. <sup>b</sup> Multiplier used to adjust all death counts to the July 1, 2019, population of 307,657,969.

**Appendix 3.** Example Reporting Lag Calculations by Category^a^

| Weeks lag<sup>b</sup> | Mean (SD) Percentage Change in Reported Deaths<br>from Earlier to Later One-Week Reporting Increment |  |  |  |  |
| --- | --- | --- | --- | --- | --- |
|  | All-Causes | Natural Causes | Heart Disease | Cancer | Chronic LRD |
| 1.6 | 32.0 (8.2) | 30.6 (8.2) | 29.2 (8.6) | 27.3 (8.6) | 27.3 (8.2) |
| 2.6 | 8.9 (2.6) | 8.4 (2.6) | 8.8 (2.7) | 6.8 (2.2) | 8.2 (2.7) |
| 3.6 | 3.3 (0.6) | 2.9 (0.5) | 3.7 (0.6) | 2.3 (0.5) | 3.1 (0.4) |
| 4.6 | 1.7 (0.3) | 1.3 (0.2) | 2.0 (0.3) | 1.1 (0.2) | 1.7 (0.5) |
| 5.6 | 1.0 (0.2) | 0.7 (0.2) | 1.4 (0.2) | 0.6 (0.1) | 1.0 (0.3) |
| 6.6 | 0.6 (0.1) | 0.3 (0.1) | 0.9 (0.2) | 0.3 (0.1) | 0.5 (0.2) |
| 7.6 | 0.4 (0.1) | 0.2 (0.1) | 0.7 (0.1) | 0.3 (0.1) | 0.3 (0.1) |
| 8.6 | 0.3 (0.1) | 0.1 (0.1) | 0.5 (0.1) | 0.2 (0.0) | 0.2 (0.1) |
| 9.6 | 0.2 (0.1) | 0.1 (0.1) | 0.4 (0.1) | 0.2 (0.1) | 0.2 (0.1) |
| 10.6 | 0.2 (0.0) | 0.0 (0.1) | 0.3 (0.1) | 0.2 (0.1) | 0.2 (0.1) |
| 11.6 | 0.1 (0.0) | 0.0 (0.1) | 0.3 (0.1) | 0.1 (0.0) | 0.2 (0.1) |
| 12.6 | 0.1 (0.1) | 0.0 (0.1) | 0.3 (0.1) | 0.1 (0.1) | 0.2 (0.1) |
| 13.6 | 0.1 (0.2) | 0.0 (0.2) | 0.3 (0.2) | 0.1 (0.2) | 0.1 (0.2) |
| 14.6 | 0.2 (0.3) | 0.1 (0.3) | 0.3 (0.3) | 0.1 (0.3) | 0.2 (0.4) |
| 15.6 | 0.1 (0.1) | 0.0 (0.2) | 0.2 (0.2) | 0.1 (0.1) | 0.1 (0.2) |
| 16.6 | 0.1 (0.1) | 0.0 (0.1) | 0.2 (0.1) | 0.1 (0.0) | 0.1 (0.1) |
| Compounded adjustment multipliers using mean values (shown through 8.6 weeks)<sup>c</sup> |  |  |  |  |  |
| 2.6<sup>b</sup> | 1.183 | 1.146 | 1.217 | 1.128 | 1.172 |
| 3.6 | 1.086 | 1.058 | 1.119 | 1.057 | 1.083 |
| 4.6 | 1.051 | 1.028 | 1.078 | 1.033 | 1.051 |
| 5.6 | 1.034 | 1.014 | 1.057 | 1.022 | 1.034 |
| 6.6 | 1.024 | 1.008 | 1.043 | 1.016 | 1.024 |
| 7.6 | 1.018 | 1.005 | 1.034 | 1.013 | 1.018 |
| 8.6 | 1.014 | 1.003 | 1.027 | 1.010 | 1.015 |
| Compounded adjustment multipliers using maximum values (shown through 8.6 weeks)<sup>c</sup> |  |  |  |  |  |
| 2.6<sup>b</sup> | 1.261 | 1.230 | 1.238 | 1.202 | 1.279 |
| 3.6 | 1.118 | 1.094 | 1.113 | 1.089 | 1.128 |
| 4.6 | 1.075 | 1.055 | 1.074 | 1.056 | 1.088 |
| 5.6 | 1.054 | 1.038 | 1.054 | 1.042 | 1.064 |
| 6.6 | 1.042 | 1.029 | 1.046 | 1.034 | 1.051 |
| 7.6 | 1.035 | 1.025 | 1.040 | 1.029 | 1.044 |
| 8.6 | 1.030 | 1.022 | 1.035 | 1.025 | 1.038 |

<sup>a</sup> Although only all-cause, natural-cause, and top 3 causes of death are shown here, all calculations were performed separately for all diagnostic categories. Each lag calculation is based on 6-7 combinations of reporting week/update week combinations beginning with reporting week ending February 1, 2020 and ending June 20, 2020, for 7 weekly updates beginning May 27, 2020, and ending July 8, 2020. <sup>b</sup> Weeks elapsed from end of reporting week to date of update. Because one state reported no deaths during the week ending June 27, its data could not be used; therefore, the smallest lag time in the file was 2.6 weeks (from week end June 20 to reporting date July 8). <sup>c</sup> Although multipliers are shown only through 8.6 weeks, adjustments were made through 16.6-week lags. SD-standard deviation.

**Appendix 4.**
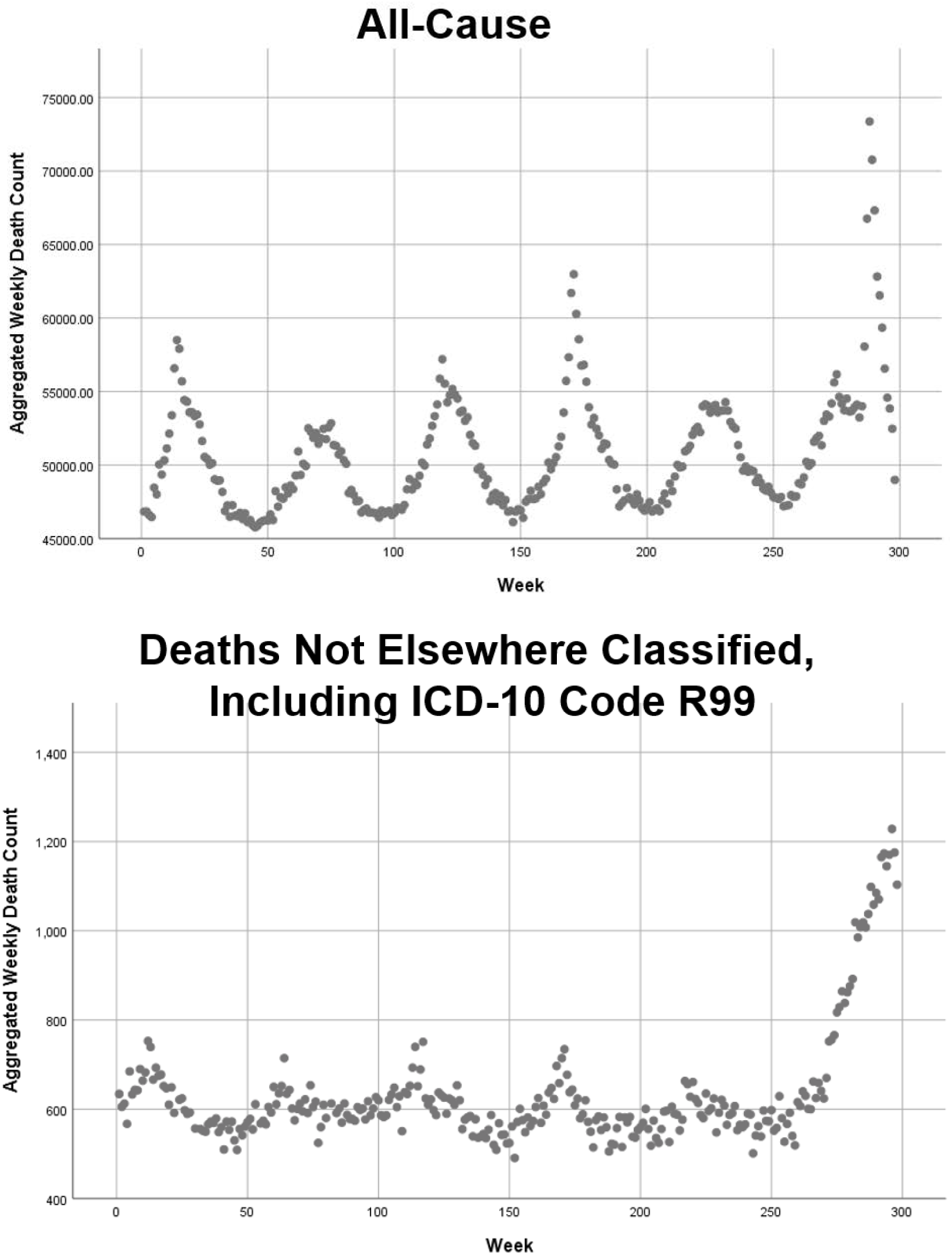
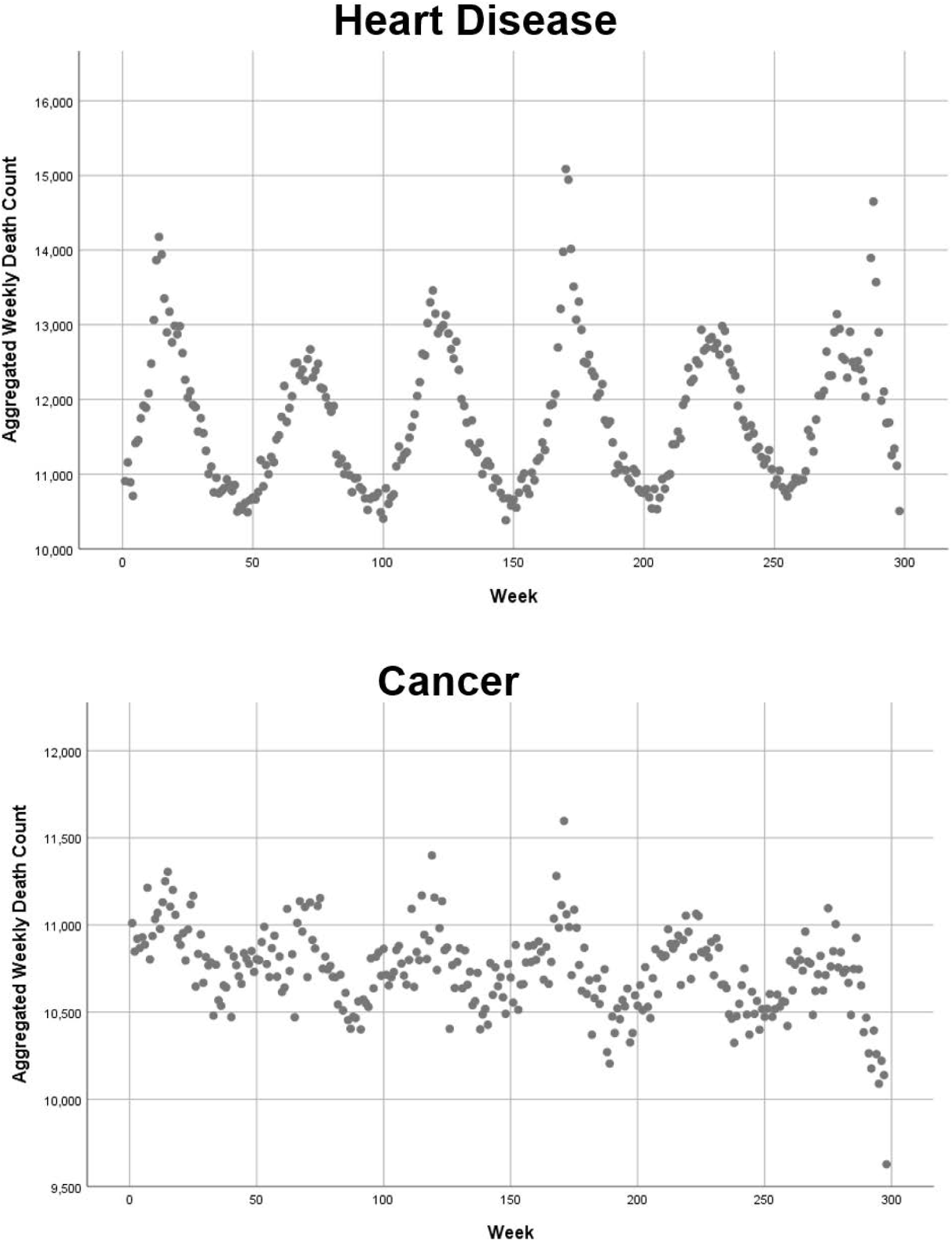
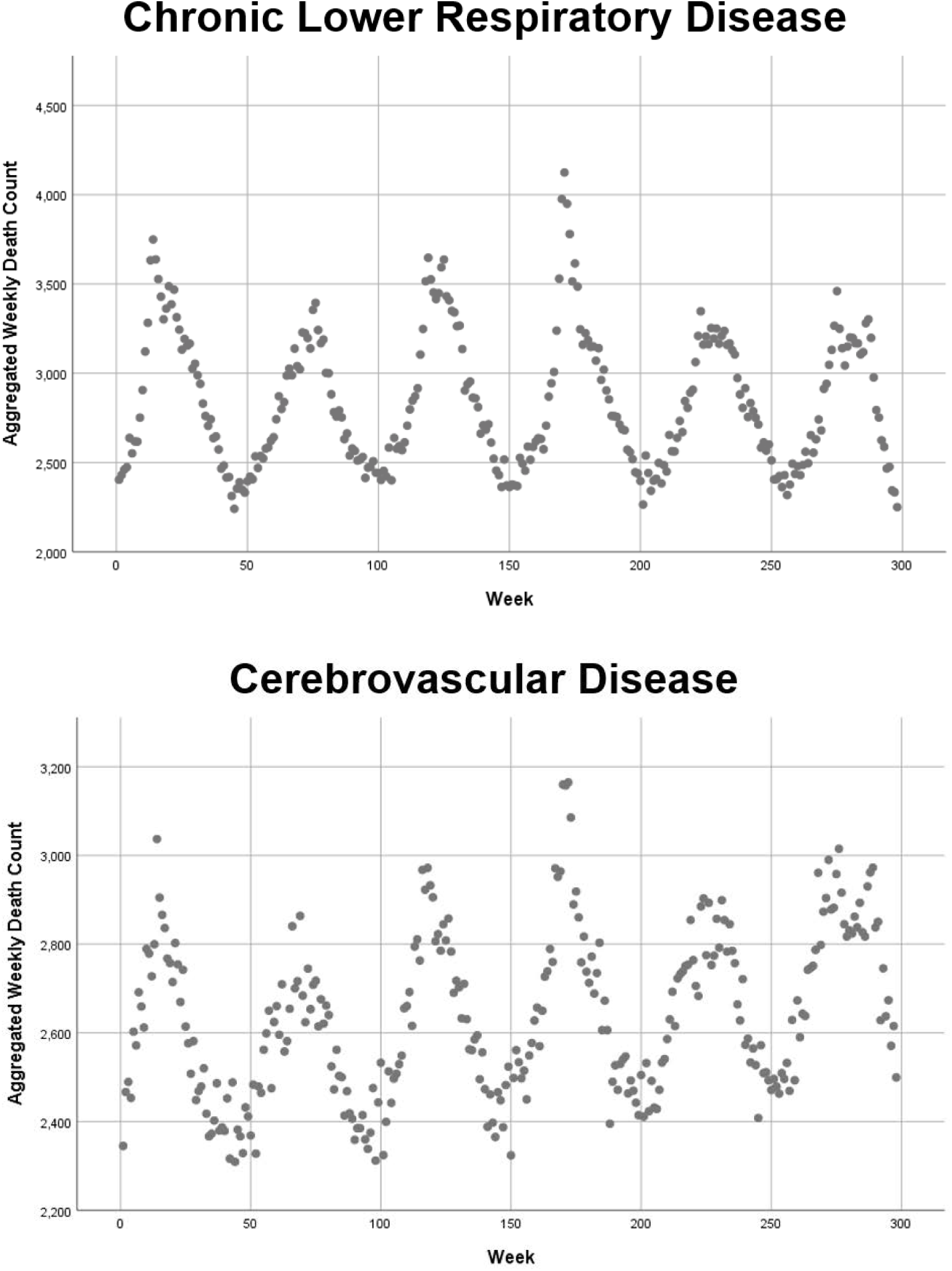
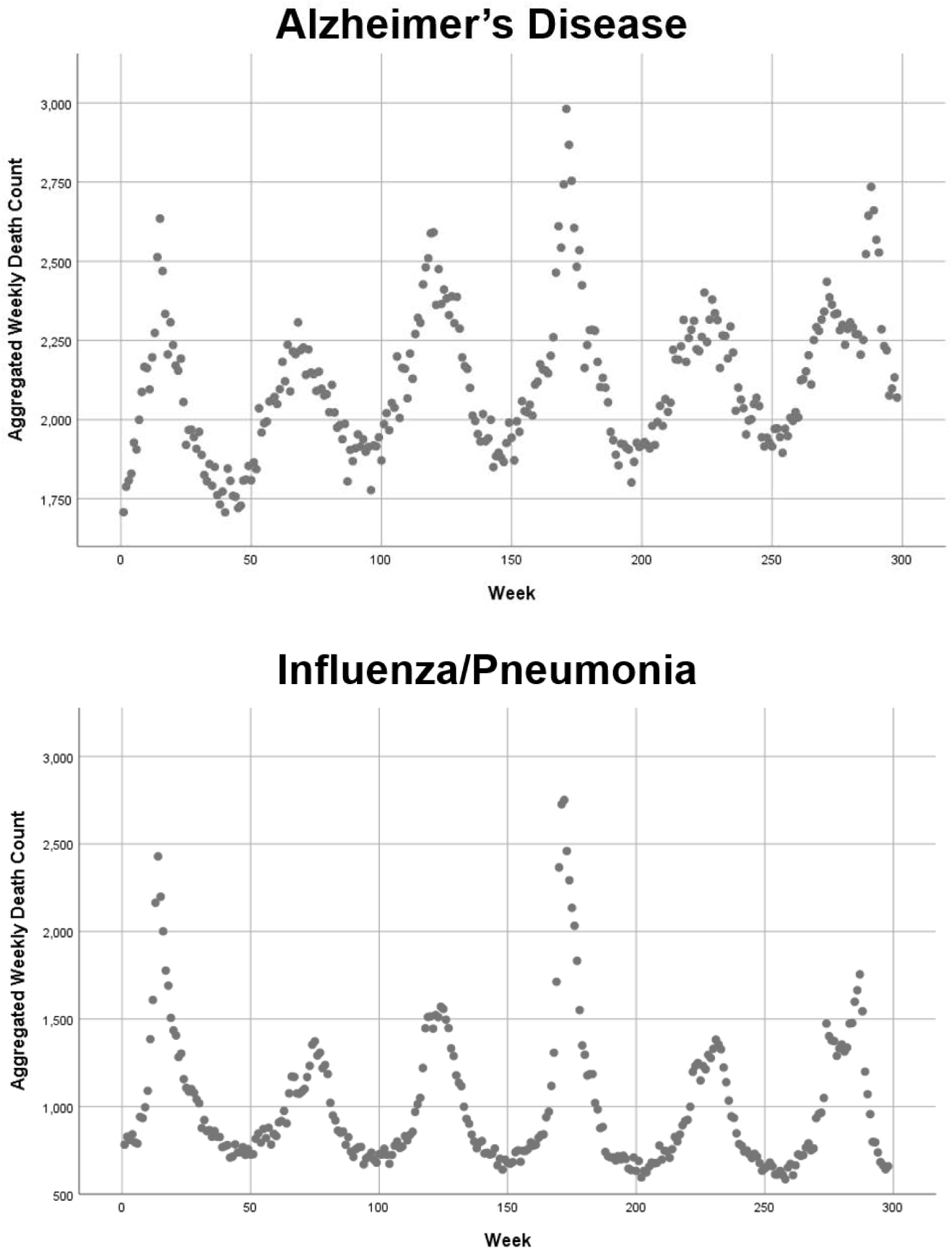
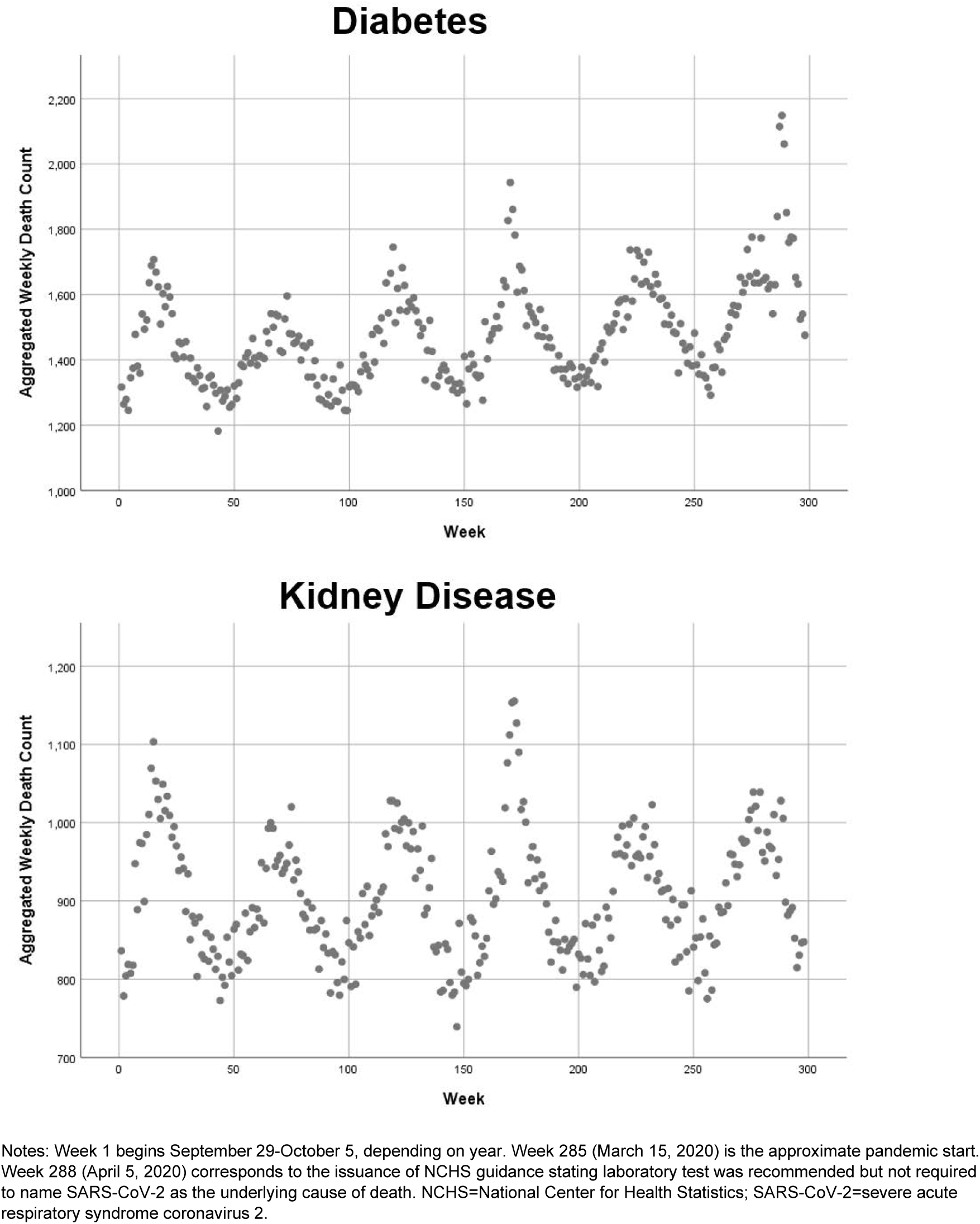
Population-Adjusted and Lag-Adjusted Weekly Death Counts, All-Cause, Not Elsewhere Classified, and Top 8 Natural Causes of Death, October 2014 through June 20, 2020, Reported as of July 8, 2020

**Appendix 5.**
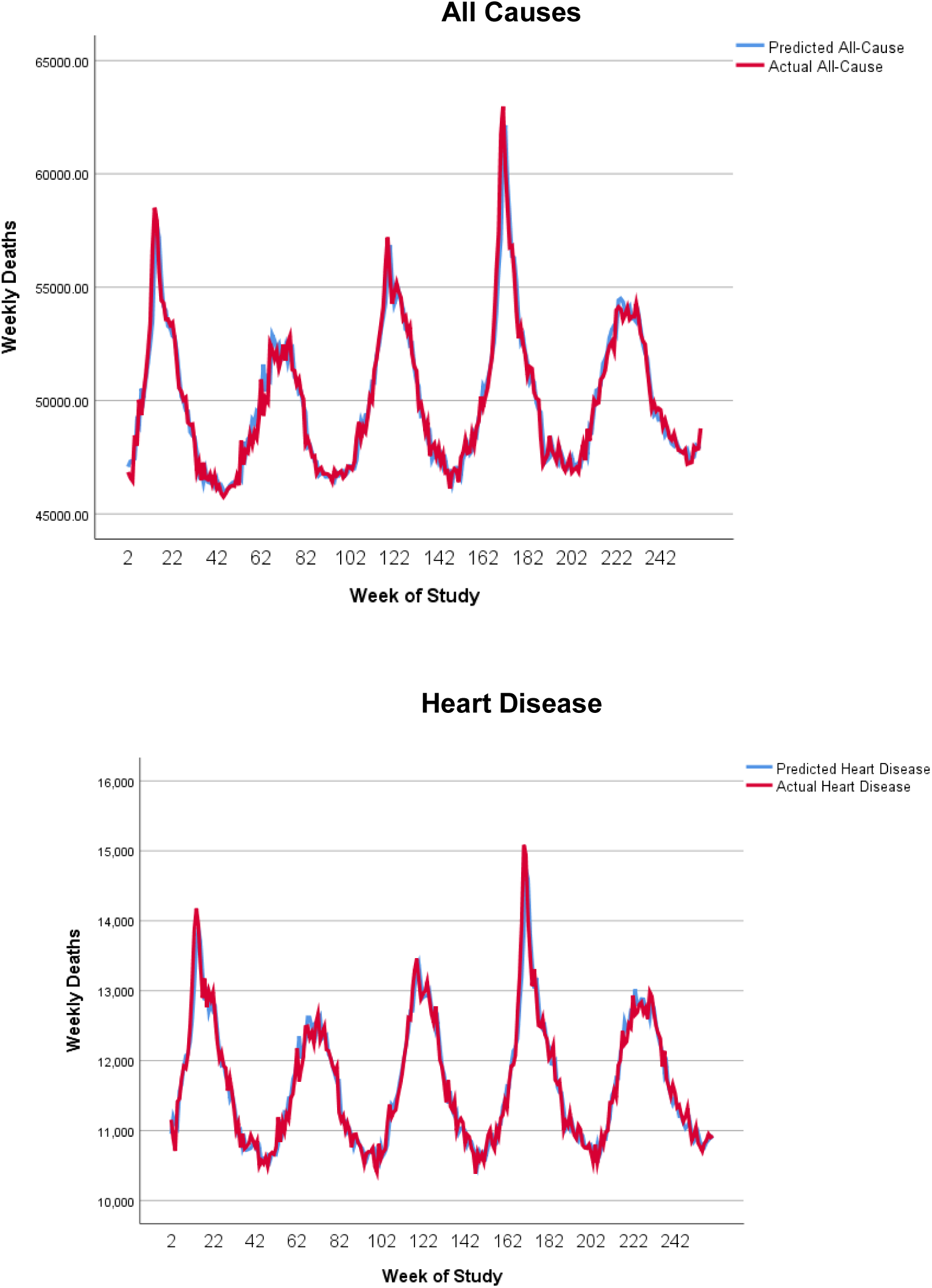
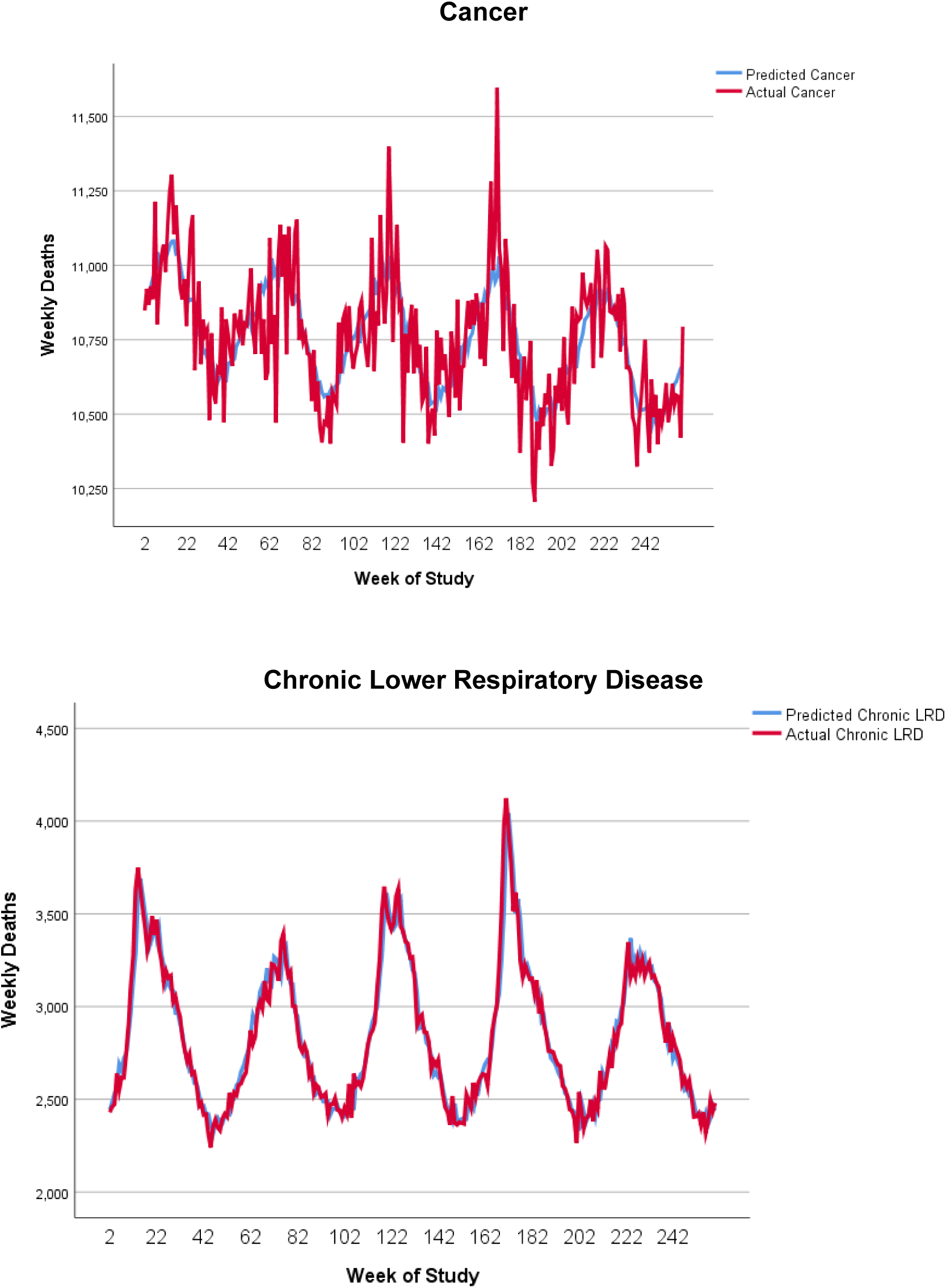
Time Series Model-Predicted Versus Actual Weekly Deaths, Calibration Period, October 2014 through September 2019, Deaths from All Causes and Top 3 Specific Natural Causes

**Appendix 6.** Summary of Projected Versus Actual Population-Adjusted Death Counts by Pandemic Time Period, Reported Through June 20 as of July 8, 2020, Time Series Analysis

|  | Actual | Projected, Using First-Order Autoregression |  |  |  | Projected, Allowing Violation of Nonautocorrelation Assumption |  |  |  |
| --- | --- | --- | --- | --- | --- | --- | --- | --- | --- |
|  |  | Point Estimate | 95% CI Lower | 95% CI Upper | Difference | Point Estimate | 95% CI Lower | 95% CI Upper | Difference |
| *Pandemic awareness (Jan 19-Mar 14)* |  |  |  |  |  |  |  |  |  |
| All-cause | 431,021 | 433,187 | 413,231 | 454,356 | -2,166 | 440,203 | 433,804 | 446,427 | -9,182 |
| Heart disease | 99,822 | 100,860 | 92,064 | 110,497 | -1,038 | 103,615 | 100,686 | 106,539 | -3,793 |
| Cancer | 86,076 | 86,098 | 71,228 | 104,066 | -22 | 86,076 | 86,086 | 83,555 | 88,637 |
| CLRD | 25,171 | 25,707 | 22,790 | 28,839 | -536 | 26,839 | 25,303 | 28,448 | -1,668 |
| NEC<sup>a</sup> | 7,344 | 4,892 | 4,310 | 5,551 | 2,452 | 4,892 | 4,310 | 5,551 | 2,452 |
| Total QM<sup>b</sup> | 117,018 | 117,984 | 104,259 | 133,520 | -966 | 118,862 | 116,000 | 122,010 | -1,844 |
| Total some QM<sup>b</sup> | 133,600 | 134,296 | 126,033 | 143,168 | -696 | 137,916 | 134,373 | 141,185 | -4,316 |
| Total non-QM<sup>b</sup> | 46,432 | 47,055 | 41,806 | 53,226 | -623 | 48,478 | 46,450 | 50,616 | -2,046 |
| *Pandemic height (Mar 15-May 16)* |  |  |  |  |  |  |  |  |  |
| All-cause | 453,098 | 431,265 | 410,994 | 452,831 | 21,833 | 365,155 | 360,482 | 369,443 | 87,943 |
| Heart disease | 91,663 | 89,345 | 81,864 | 97,511 | 2,318 | 84,971 | 82,912 | 87,003 | 6,692 |
| Cancer | 74,190 | 74,053 | 61,644 | 88,954 | 137 | 74,038 | 72,161 | 75,887 | 152 |
| CLRD | 21,420 | 21,503 | 19,278 | 23,845 | -83 | 21,796 | 20,731 | 22,890 | -376 |
| NEC<sup>a</sup> | 7,377 | 3,869 | 3,477 | 4,309 | 3,508 | 7,377 | 3,869 | 3,477 | 4,309 |
| Total QM<sup>b</sup> | 100,937 | 100,822 | 89,609 | 113,449 | 115 | 100,673 | 98,668 | 102,918 | 264 |
| Total some QM<sup>b</sup> | 121,649 | 118,886 | 111,974 | 126,223 | 2,763 | 111,679 | 109,255 | 113,985 | 9,970 |
| Total non-QM<sup>b</sup> | 44,462 | 42,426 | 37,874 | 47,784 | 2,036 | 39,258 | 37,860 | 40,704 | 5,204 |
| *Late pandemic (May 17-June 20)* |  |  |  |  |  |  |  |  |  |
| All-cause | 387,338 | 387,489 | 370,653 | 405,326 | -151 | 344,374 | 340,345 | 348,142 | 42,964 |
| Heart disease | 79,692 | 80,005 | 73,870 | 86,650 | -313 | 79,143 | 77,368 | 80,897 | 549 |
| Cancer | 70,906 | 72,890 | 61,003 | 87,089 | -1,984 | 73,160 | 71,435 | 74,860 | -2,254 |
| CLRD | 17,082 | 17,915 | 16,282 | 19,606 | -833 | 19,097 | 18,231 | 19,976 | -2,015 |
| NEC<sup>a</sup> | 8,160 | 3,828 | 3,468 | 4,231 | 4,332 | 3,828 | 3,468 | 4,231 | 4,332 |
| Total QM<sup>b</sup> | 92,602 | 94,880 | 84,852 | 106,084 | -2,278 | 96,700 | 94,924 | 98,653 | -4,098 |
| Total some QM<sup>b</sup> | 103,039 | 103,526 | 98,119 | 109,254 | -487 | 102,376 | 100,362 | 104,430 | 663 |
| Total non-QM<sup>b</sup> | 37,464 | 37,286 | 33,645 | 41,510 | 178 | 36,095 | 34,904 | 37,313 | 1,369 |

<sup>a</sup> QM diagnoses are cancer, CLRD, and septicemia. Some QM (56%-81% of reported category) diagnoses are cerebrovascular disease, heart disease, and influenza/pneumonia. No QM diagnoses are Alzheimer's disease, diabetes, and other (mostly acute) respiratory infections. CI=confidence interval; CLRD=chronic lower respiratory disease; NEC=not elsewhere classified; QM=quality metric.

## Data Availability

Data used in this study are publicly available. Details of study data files available upon reasonable request.

